# SARS-CoV-2 Omicron has extensive but incomplete escape of Pfizer BNT162b2 elicited neutralization and requires ACE2 for infection

**DOI:** 10.1101/2021.12.08.21267417

**Authors:** Sandile Cele, Laurelle Jackson, David S. Khoury, Khadija Khan, Thandeka Moyo-Gwete, Houriiyah Tegally, James Emmanuel San, Deborah Cromer, Cathrine Scheepers, Daniel Amoako, Farina Karim, Mallory Bernstein, Gila Lustig, Derseree Archary, Muneerah Smith, Yashica Ganga, Zesuliwe Jule, Kajal Reedoy, Shi-Hsia Hwa, Jennifer Giandhari, Jonathan M. Blackburn, Bernadett I. Gosnell, Salim S. Abdool Karim, Willem Hanekom, NGS-SA, COMMIT-KZN Team, Anne von Gottberg, Jinal Bhiman, Richard J. Lessells, Mahomed-Yunus S. Moosa, Miles P. Davenport, Tulio de Oliveira, Penny L. Moore, Alex Sigal

## Abstract

The emergence of SARS-CoV-2 Omicron, first identified in Botswana and South Africa, may compromise vaccine effectiveness and the ability of antibodies triggered by previous infection to protect against re-infection (1). Here we investigated whether Omicron escapes antibody neutralization in South Africans, either previously SARS-CoV-2 infected or uninfected, who were vaccinated with Pfizer BNT162b2. We also investigated if Omicron requires the ACE2 receptor to infect cells. We isolated and sequence confirmed live Omicron virus from an infected person in South Africa and compared plasma neutralization of this virus relative to an ancestral SARS-CoV-2 strain with the D614G mutation, observing that Omicron still required ACE2 to infect. For neutralization, blood samples were taken soon after vaccination, so that vaccine elicited neutralization was close to peak. Neutralization capacity of the D614G virus was much higher in infected and vaccinated versus vaccinated only participants but both groups had 22-fold Omicron escape from vaccine elicited neutralization. Previously infected and vaccinated individuals had residual neutralization predicted to confer 73% protection from symptomatic Omicron infection, while those without previous infection were predicted to retain only about 35%. Both groups were predicted to have substantial protection from severe disease. These data support the notion that high neutralization capacity elicited by a combination of infection and vaccination, and possibly boosting, could maintain reasonable effectiveness against Omicron. A waning neutralization response is likely to decrease vaccine effectiveness below these estimates. However, since protection from severe disease requires lower neutralization levels and involves T cell immunity, such protection may be maintained.

---

The emergence of the Omicron variant (https://www.nicd.ac.za/wp-content/uploads/2021/11/Update-of-SA-sequencing-data-from-GISAID-26-Nov_Final.pdf) of SARS-CoV-2 in November 2021 in South Africa and Botswana has raised concerns that, based on the large number of mutations in the spike protein and elsewhere on the virus (https://covdb.stanford.edu/page/mutation-viewer/#omicron), this variant will have considerable escape from vaccine elicited immunity. Furthermore, several mutations in the receptor binding domain and S2 are predicted to impact transmissibility and affinity for human ACE2 (hACE2).

We previously engineered a human lung cell line (H1299-ACE2, Fig S1) to over-express the hACE2 receptor (2). Here we used it here to both isolate Omicron and test neutralization (Materials and methods). Isolation of the Omicron virus was done using two passages in H1299-ACE2, with the second passage a coculture of infected H1299-ACE2 with the Vero E6 African green monkey kidney cell line. Sequencing of the isolated virus confirmed it was the Omicron variant bearing the R346K mutation. There were no in vitro introduced mutations as majority or minority variants (Table S1). H1299-ACE2 cells were similar to Vero E6 in the formation of infection foci in a live virus infection with ancestral D614G and Beta variant viruses but were considerably more sensitive relative to unmodified Vero E6 (Fig S2A-B). Infection by cell-free Omicron of unmodified Vero E6 cells was poor (Fig S2C) and we were unable to use cell-free Omicron infection in Vero E6 cells to generate a useable virus stock (Fig S2D).

We first tested the isolated virus on the H1299-ACE2 and H1299 parental cells and observed that Omicron infected the hACE2-expressing cells in a concentration dependent manner but did not infect the parental H1299, indicating that hACE2 is required for Omicron entry (Fig. 1A-B). We then tested the ability of plasma from BNT162b2 vaccinated study participants to neutralize Omicron versus ancestral D614G virus in a live virus neutralization assay. We tested plasma samples from 19 participants (Table S2, S3), with 6 having no previous record of SARS-CoV-2 infection nor detectable SARS-CoV-2 nucleocapsid antibodies indicative of previous infection. Samples from a later timepoint were available for two of the vaccinated only participants (Table S3) and these were also tested. The previously infected and vaccinated participants were infected with either ancestral strains or the Delta variant (Table S3). To quantify neutralization in the live virus neutralization assay, we calculated the focus reduction neutralization test (FRNT_50_) value, which is the inverse of the plasma dilution required for 50% reduction in infection focus number.

**Figure 1:**
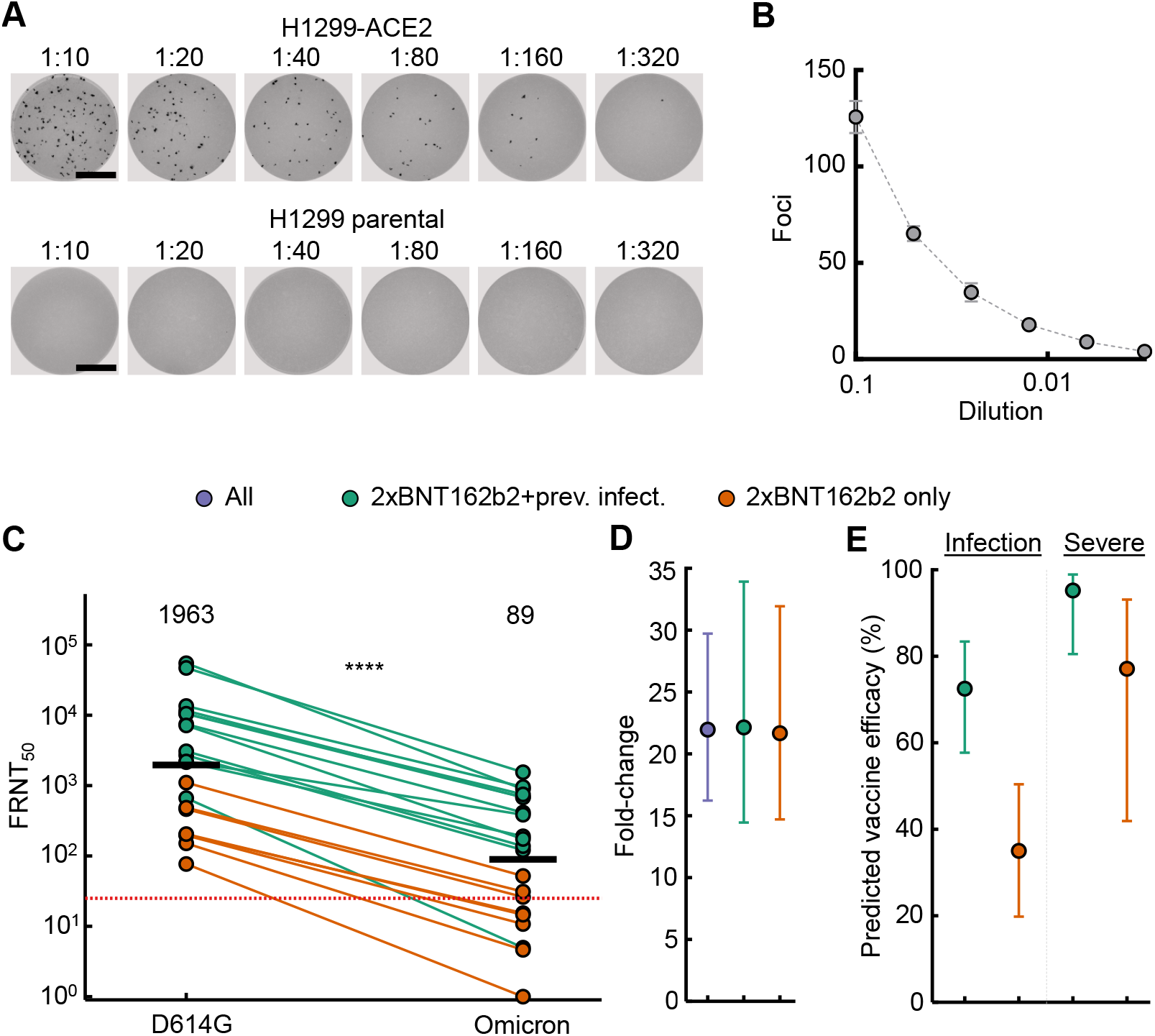
ACE2 dependence and neutralization of the Omicron variant by Pfizer BNT162b2 elicited immunity. (A) Images of infection foci in titration of live SARS-CoV-2 Omicron virus on H1299-ACE2 and H1299 parental cells. Numbers above well images denote viral stock dilution. Scale bar is 2mm. (B) Quantified number of foci as a function of Omicron virus stock dilution. Mean and standard deviation of 6 replicates from 2 independent experiments. (C) Neutralization of Omicron virus compared to D614G ancestral virus by plasma from participants vaccinated with BNT162b2 and previously SARS-CoV-2 infected (green) or vaccinated only (orange). Numbers in black above each virus strain are geometric mean titers (GMT) of the reciprocal plasma dilution (FRNT50) resulting in 50% reduction in the number of infection foci. Red horizontal line denotes most concentrated plasma used. 21 samples were tested from 19 participants in 2 independent experiments, where n=6 were vaccinated only and n=13 were vaccinated and previously infected. p=4.8 *×*10^*−*5^ as determined by the Wilcoxon rank sum test. (D) Geometric means and 95% confidence intervals of fold-changes between participant FRNT50 values for ancestral D614G versus Omicron neutralization. Purple denotes all participants, green denotes vaccinated previously infected and orange denotes vaccinated only. (E) Prediction of vaccine efficacy against symptomatic infection (left) and severe disease (right) based on Omicron neutralization results.

Consistent with previous studies (3-5), we observed that previously infected and vaccinated individuals had higher neutralization capacity of ancestral virus relative to vaccinated only participants (Fig 1C). For all participants, the ability to neutralize Omicron was lower than ancestral virus (Fig 1C). Geometric mean titer (GMT) FRNT_50_ for all participants declined from 1963 to 89, a 22-fold drop (Fig 1D, 95% CI 16-30). The fold drop was 22-fold both in individuals who were previously infected and vaccinated (95% CI 16-34) and vaccinated only (95% CI 15-32) despite the higher neutralization capacity of the vaccinated and previously infected participants (Fig 1D). We compared these results with neutralization of the Beta variant (2, 6-12) using Beta and ancestral virus infection of H1299-ACE2 (Fig S3A) and Vero E6 (Fig S3B) cells. Fold-drop relative to the ancestral D614G virus was 4.3 for H1299-ACE2 and 5.0 for Vero E6. These two cell lines therefore gave similar results and showed approximately 4 to 5-fold greater escape of Omicron relative to Beta.

The higher residual Omicron neutralization capacity in previously infected vaccinated participants is expected to translate to higher vaccine efficacy (13, 14). We therefore calculated the predicted vaccine efficacy for protecting against symptomatic infection and severe disease using a previously developed model which fits neutralization values to vaccine efficacy data from randomized control trials (13, 14). A limitation of this approach for predicting vaccine efficacy against severe disease is that, although a relationship between neutralization and protection from severe outcomes has been demonstrated, there is considerably more uncertainty in this estimate given lower numbers of severe cases in the randomized control trials. Furthermore, protection against severe outcomes may vary depending on how the pathogenicity of Omicron infections compares to other SARS-CoV-2 variants. Using this model, we estimate a vaccine efficacy for preventing Omicron symptomatic infection of 73% (95% CI 58-83%) for vaccinated and previously infected individuals and 35% (95% CI 20-50%) for vaccinated only participants, essentially compromising the ability of the vaccine to protect against infection in the latter group (Fig 1E). Efficacy against severe disease is predicted to be 95% (95% CI 81-99%) for vaccinated and previously infected individuals and 77% (95% CI 42-93%) for vaccinated only (Fig 1E). For the vaccinated only group, confidence intervals are wide but indicate substantial protection.

Shortly after we released results, several other groups reported results (15) including Pfizer-BioNTech (https://www.businesswire.com/news/home/20211208005542/en/). These results mirror ours, with large fold-drops in neutralization of Omicron versus ancestral virus. Interestingly, the Pfizer-BioNTech study reports that boosting greatly reduces Omicron escape. We do not see such a qualitative effect in the vaccinated previously infected participants in this study and instead we observe that samples with high neutralization have very similar fold-drops to samples with lower neutralization.

Limitations of this study include the inaccuracy inherent in predicting protection from severe disease. Also, the R346K substitution in the isolate, a putative escape mutation (16) which may confer additional antibody resistance (https://jbloomlab.github.io/SARS2_RBD_Ab_escape_maps/escape-calc/), is not found in the majority of Omicron genomes. Importantly, the timing of sample collection soon after vaccination (Table S1, S2) does not account for the waning of neutralization capacity (17, 18) which will reduce vaccine efficacy estimates. However, while T cell immunity against Omicron has yet to be measured, such responses have thus far maintained activity against Beta and Delta, and will likely contribute to protection from severe disease caused by Omicron infection (19, 20) and may counterbalance the effects of waning neutralization. While there may be other explanations for lower pathogenicity in an emerging variant (21), the incomplete escape of Omicron from neutralization may predict that the frequency of severe disease in an Omicron infection wave would be less than in previous infection waves where immunity to SARS-CoV-2 at the population level was lower.

## Materials and methods

### Ethical statement

Blood samples were obtained from hospitalized adults with PCR-confirmed SARS-CoV-2 infection and/or vaccinated individuals who were enrolled in a prospective cohort study approved by the Biomedical Research Ethics Committee at the University of KwaZulu–Natal (reference BREC/00001275/2020). Use of residual swab sample was approved by the University of the Witwatersrand Human Research Ethics Committee (HREC) (ref. M210752).

### Whole-genome sequencing, genome assembly and phylogenetic analysis

cDNA synthesis was performed on the extracted RNA using random primers followed by gene-specific multiplex PCR using the ARTIC V.3 protocol (https://www.protocols.io/view/covid-19-artic-v3-illumina-library-construction-an-bibtkann). In brief, extracted RNA was converted to cDNA using the Superscript IV First Strand synthesis system (Life Technologies) and random hexamer primers. SARS-CoV-2 whole-genome amplification was performed by multiplex PCR using primers designed using Primal Scheme (http://primal.zibraproject.org/) to generate 400-bp amplicons with an overlap of 70 bp that covers the 30 kb SARS-CoV-2 genome. PCR products were cleaned up using AmpureXP purification beads (Beckman Coulter) and quantified using the Qubit dsDNA High Sensitivity assay on the Qubit 4.0 instrument (Life Technologies). We then used the Illumina Nextera Flex DNA Library Prep kit according to the manufacturer’s protocol to prepare indexed paired-end libraries of genomic DNA. Sequencing libraries were normalized to 4 nM, pooled and denatured with 0.2 N sodium acetate. Then, a 12-pM sample library was spiked with 1% PhiX (a PhiX Control v.3 adaptor-ligated library was used as a control). We sequenced libraries on a 500-cycle v.2 MiSeq Reagent Kit on the Illumina MiSeq instrument (Illumina). We assembled paired-end fastq reads using Genome Detective 1.126 (https://www.genomedetective.com) and the Coronavirus Typing Tool. We polished the initial assembly obtained from Genome Detective by aligning mapped reads to the reference sequences and filtering out low-quality mutations using the bcftools 1.7-2 mpileup method. Mutations were confirmed visually with BAM files using Geneious software (Biomatters). P2 stock was sequenced and confirmed Omicron with the following substitutions: E:T9I,M:D3G,M:Q19E,M:A63T,N:P13L,N:R203K,N:G204R,ORF1a:K856R,ORF1a:L2084I,ORF1a:A2710T, ORF1a:T3255I,ORF1a:P3395H,ORF1a:I3758V,ORF1b:P314L,ORF1b:I1566V,ORF9b:P10S,S:A67V,S:T95I ,S:Y145D,S:L212I,S:G339D,S:R346K,S:S371L,S:S373P,S:S375F,S:K417N,S:N440K,S:G446S,S:S477N,S:T4 78K,S:E484A,S:Q493R,S:G496S,S:Q498R,S:N501Y,S:Y505H,S:T547K,S:D614G,S:H655Y,S:N679K,S:P681 H,S:N764K,S:D796Y,S:N856K,S:Q954H,S:N969K,S:L981F. Deletions: N:E31-,N:R32-,N:S33-,ORF1a:S2083-,ORF1a:L3674-,ORF1a:S3675-,ORF1a:G3676-,ORF9b:E27-,ORF9b:N28-,ORF9b:A29-,S:H69-,S:V70-,S:G142-,S:V143-,S:Y144-,S:N211-.Sequence was deposited in GISAID, accession: EPI_ISL_7358094.

### SARS-CoV-2 nucleocapsid enzyme-linked immunosorbent assay (ELISA)

2 μg/ml nucleocapsid protein (Biotech Africa; Catalogue number: BA25-P was used to coat 96-well, high-binding plates and incubated overnight at 4°C. The plates were incubated in a blocking buffer consisting of 5% skimmed milk powder, 0.05% Tween 20, 1x PBS. Plasma samples were diluted to a 1:100 dilution in a blocking buffer and added to the plates. IgG secondary antibody was diluted to 1:3000 in blocking buffer and added to the plates followed by TMB substrate (Thermo Fisher Scientific). Upon stopping the reaction with 1 M H2SO4, absorbance was measured at a 450 nm wavelength.

### Cells

Vero E6 cells (ATCC CRL-1586, obtained from Cellonex in South Africa) were propagated in complete DMEM with 10% fetal bovine serum (Hylone) containing 1% each of HEPES, sodium pyruvate, L-glutamine and nonessential amino acids (Sigma-Aldrich). Vero E6 cells were passaged every 3–4 days. The H1299-E3 cell line for first-passage SARS-CoV-2 expansion, derived as described in (2), was propagated in complete RPMI with 10% fetal bovine serum containing 1% each of HEPES, sodium pyruvate, L-glutamine and nonessential amino acids. H1299 cells were passaged every second day. Cell lines have not been authenticated. The cell lines have been tested for mycoplasma contamination and are mycoplasma negative.

### Virus expansion

All work with live virus was performed in Biosafety Level 3 containment using protocols for SARS-CoV-2 approved by the AHRI Biosafety Committee. ACE2-expressing H1299-E3 cells were seeded at 4.5 × 10^5^ cells in a 6 well plate well and incubated for 18–20 h. After one DPBS wash, the sub-confluent cell monolayer was inoculated with 500 μL universal transport medium diluted 1:1 with growth medium filtered through a 0.45-μm filter. Cells were incubated for 1 h. Wells were then filled with 3 mL complete growth medium. After 4 days of infection (completion of passage 1 (P1)), cells were trypsinized, centrifuged at 300 rcf for 3 min and resuspended in 4 mL growth medium. Then 2 mL was added to Vero E6 cells that had been seeded at 2 × 10^5^ cells per mL, 5mL total, 18–20 h earlier in a T25 flask (approximately 1:8 donor-to-target cell dilution ratio) for cell-to-cell infection. The coculture of ACE2-expressing H1299-E3 and Vero E6 cells was incubated for 1 h and the flask was then filled with 7 mL of complete growth medium and incubated for 4 days. The viral supernatant (passage 2 (P2) stock) was used for experiments. Further optimization of the viral outgrowth protocol used for subsequent omicron isolates showed that addition of 4 mL instead of 2 mL of infected H1299-E3 cells to Vero E6 cells that had been seeded at 2 × 10^5^ cells per mL, 20 mL total, 18–20 h earlier in a T75 flask gave P2 stocks with substantially higher titers.

### Live virus neutralization assay

H1299-E3 cells were plated in a 96-well plate (Corning) at 30,000 cells per well 1 day pre-infection. Plasma was separated from EDTA-anticoagulated blood by centrifugation at 500 rcf for 10 min and stored at −80 °C. Aliquots of plasma samples were heat-inactivated at 56 °C for 30 min and clarified by centrifugation at 10,000 rcf for 5 min. Virus stocks were used at approximately 50-100 focus-forming units per microwell and added to diluted plasma. Antibody–virus mixtures were incubated for 1 h at 37 °C, 5% CO_2_. Cells were infected with 100 μL of the virus–antibody mixtures for 1 h, then 100 μL of a 1X RPMI 1640 (Sigma-Aldrich, R6504), 1.5% carboxymethylcellulose (Sigma-Aldrich, C4888) overlay was added without removing the inoculum. Cells were fixed 18 h post-infection using 4% PFA (Sigma-Aldrich) for 20 min. Foci were stained with a rabbit anti-spike monoclonal antibody (BS-R2B12, GenScript A02058) at 0.5 μg/mL in a permeabilization buffer containing 0.1% saponin (Sigma-Aldrich), 0.1% BSA (Sigma-Aldrich) and 0.05% Tween-20 (Sigma-Aldrich) in PBS. Plates were incubated with primary antibody overnight at 4 °C, then washed with wash buffer containing 0.05% Tween-20 in PBS. Secondary goat anti-rabbit horseradish peroxidase (Abcam ab205718) antibody was added at 1 μg/mL and incubated for 2 h at room temperature with shaking. TrueBlue peroxidase substrate (SeraCare 5510-0030) was then added at 50 μL per well and incubated for 20 min at room temperature. Plates were imaged in an ELISPOT instrument with built-in image analysis (C.T.L).

### Statistics and fitting

All statistics and fitting were performed using MATLAB v.2019b. Neutralization data were fit to Tx=1/1+(D/ID50).

Here Tx is the number of foci normalized to the number of foci in the absence of plasma on the same plate at dilution D and ID50 is the plasma dilution giving 50% neutralization. FRNT50 = 1/ID50. Values of FRNT50 <1 are set to 1 (undiluted), the lowest measurable value.

### Estimating vaccine efficacy from neutralization titers

Previously, the fold reduction in neutralization was shown to correlate and predict vaccine efficacy against symptomatic and severe infection with ancestral SARS-CoV-2 infection (14), and more recently with variants of concern (13). The methods used in these previous studies were used here to estimate the vaccine efficacy against Omicron based on the loss of neutralization observed in this study. Briefly, vaccine efficacy (VE) was estimated based on the (log10) mean neutralization titer as a fold of the mean convalescent titer reported for BNT162b2 in phase 1/2 trials (μ), and the (log10) fold drop in neutralization titer to Omicron (*f*) using the equation:

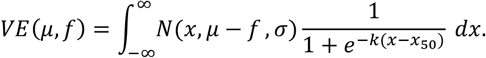

Here, *N* is the probability density function of a normal distribution with mean μ − *f* and standard deviation *σ*, and *k* and *x*_50_ are the parameters of the logistic function relating neutralization to protection. All parameters excluding *f* were estimated previously as, μ = log_10_ 2.4, *σ* = 0.46, *k* = 3 and *x*_50_ = log_10_ 0.2 for symptomatic infection and *x*_50_ = log_10_ 0.03 for severe disease (14).

## Data Availability

All data produced in the present work are contained in the manuscript

## Acknowledgements

This study was supported by the Bill and Melinda Gates award INV-018944 (AS), National Institutes of Health award R01 AI138546 (AS), and South African Medical Research Council awards (AS, TdO, PLM) and the UK Foreign, Commonwealth and Development Office and Wellcome Trust (Grant no 221003/Z/20/Z, PLM). PLM is also supported by the South African Research Chairs Initiative of the Department of Science and Innovation and the NRF (Grant No 98341). DSK, DC, and MPD are supported by NHMRC (Australia) Fellowship / Investigator grants. The funders had no role in study design, data collection and analysis, decision to publish, or preparation of the manuscript.

---

**Figure S1:**
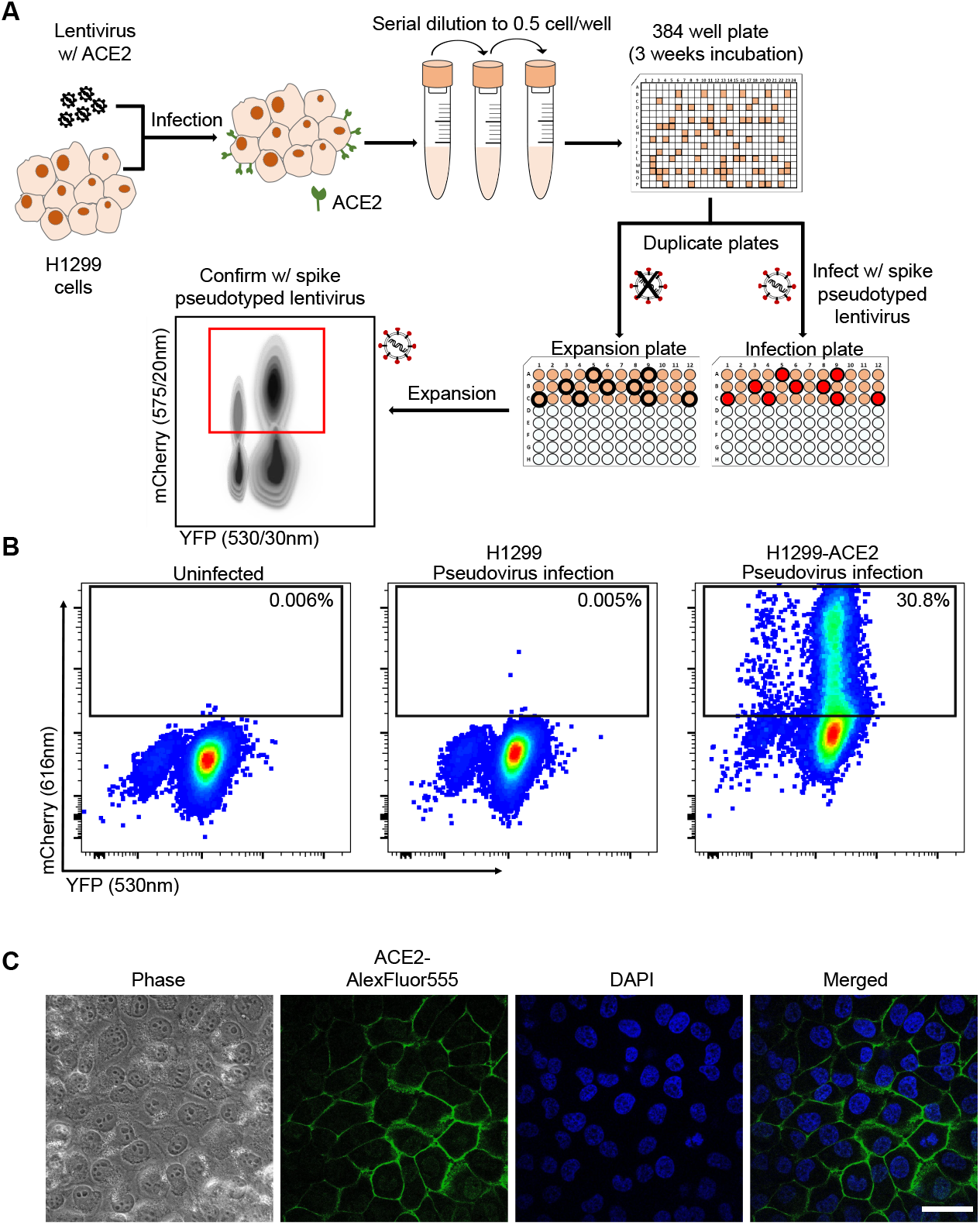
Generation of H1299-ACE2 clonal cell line. (A) The H1299 uman non-small cell lung carcinoma cell line with YFP labelled histone H2AZ was spinfected with the pHAGE2-EF1a-Int-ACE2 lentivector. Cells were single cell cloned by limiting dilution in a 384-well plate. Clones were expanded into duplicate 96-well plates, where one plate was used to select infectable clones based on mCherry signal from infection with SARS-CoV-2 mCherry expressing spike pseudotyped lentivirus. Clones were chosen based on infectability and expanded from the non-infected replicate 96-well plate. (B) Flow cytometry of SARS-CoV-2 mCherry expressing spike pseudotyped lentivirus infection in H1299-ACE2 cells. (C) Images of H1299-ACE2 cells stained with anti-ACE2 antibody and DAPI. Note membrane localization of ACE2. Scale bar is 50*µ*m.

**Figure S2:**
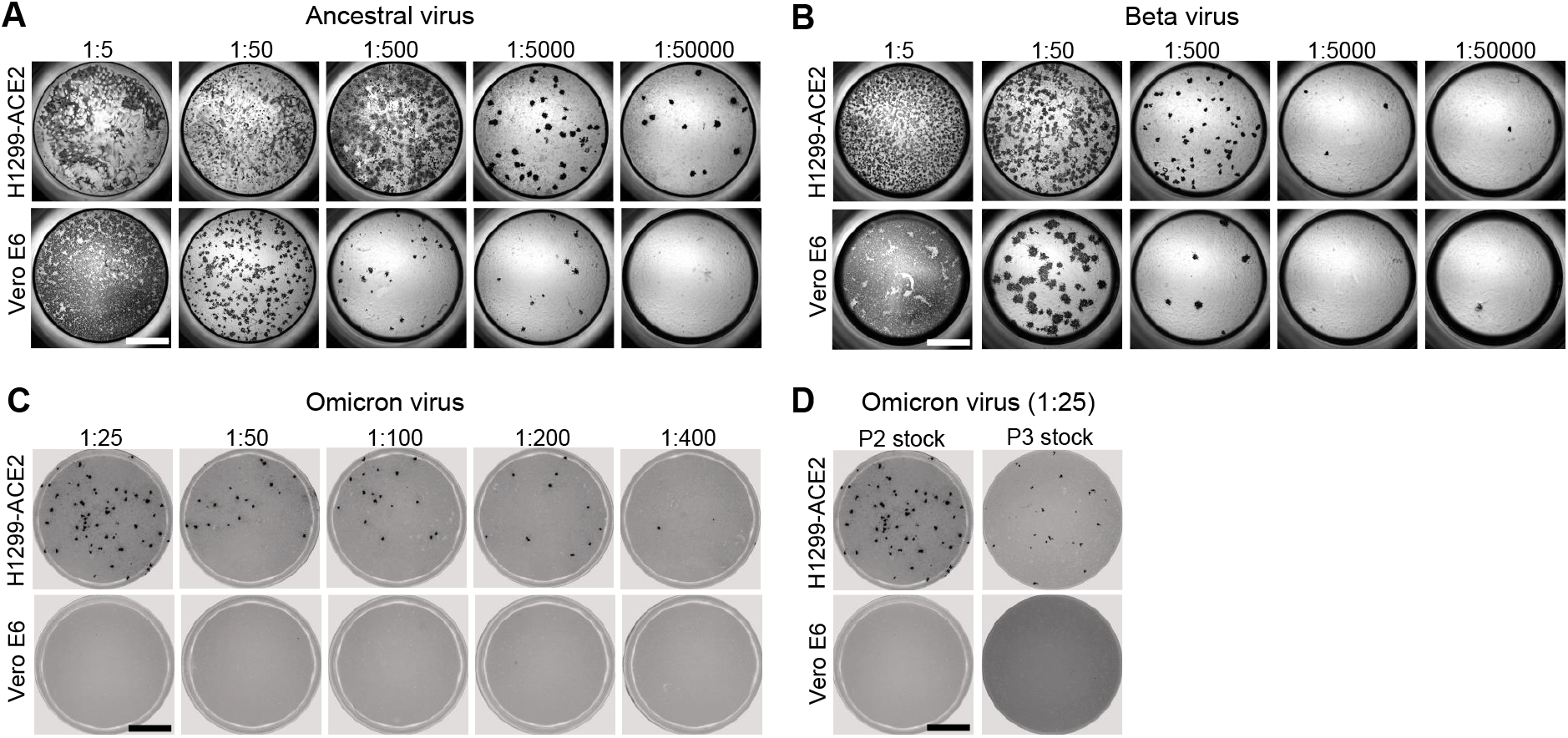
Comparison of SARS-CoV-2 infection in H1299-ACE2 and Vero E6 cells. Both H1299-ACE2 and Vero E6 cells were infected with the same viral stock in the same experiment with D614G virus (A) or Beta virus (B) and a focus forming assay was performed. (C) Focus forming assay with stock of Omicron virus isolate on H1299-ACE2 and Vero E6 cells. (D) Comparison of passage 2 (P2) and passage 3 (P3) stock, where P3 stock was generated by infection of 1 mL of cell-free P2 stock in 20 mL of Vero E6 cells seeded at 2 *×* 10^5^ cells per mL and incubated over 4 days. Numbers above well images denote viral stock dilution. Scale bar is 2mm.

**Figure S3:**
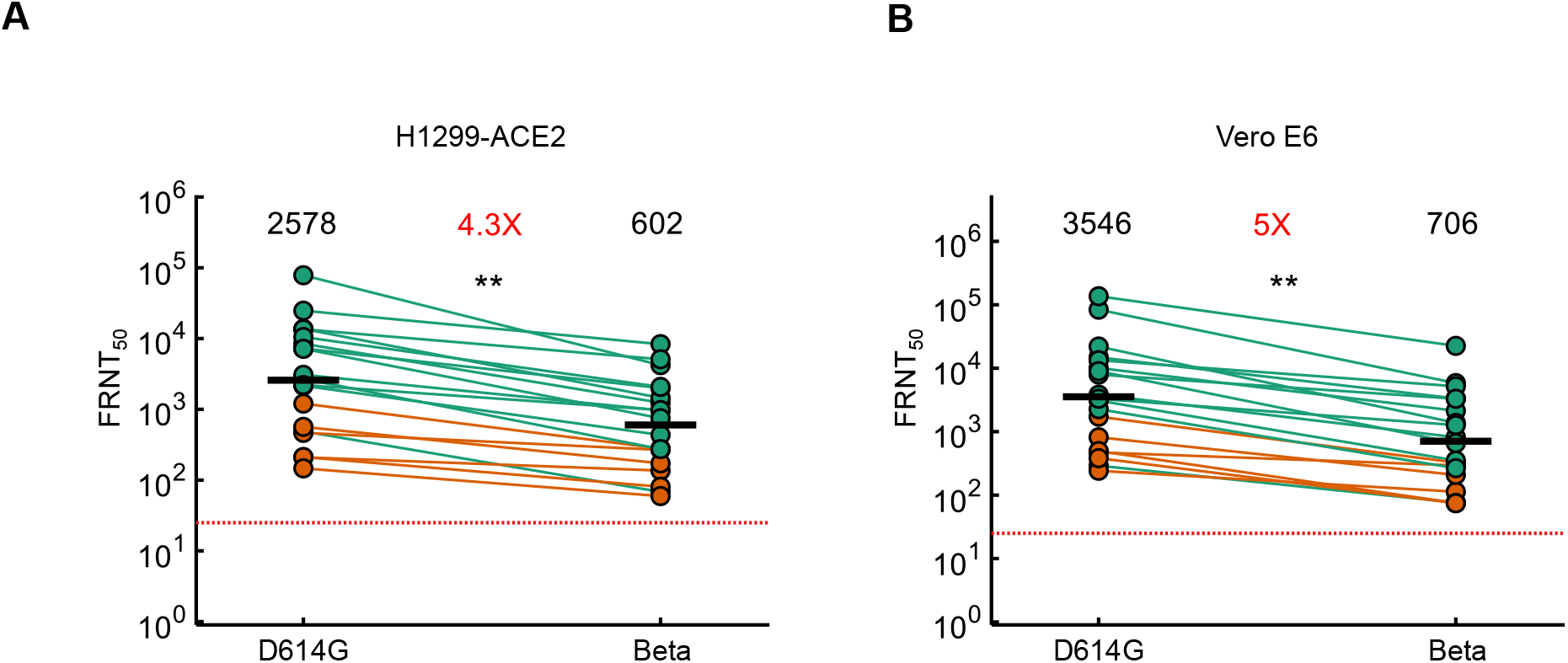
Neutralization of the Beta variant by Pfizer BNT162b2 elicited immunity. Neutralization of the Beta variant virus compared to D614G ancestral virus in H1299-ACE2 (A) or Vero E6 cells (B) in participants vaccinated with BNT162b2 and infected by SARS-CoV-2 (green) or vaccinated only (orange). Numbers in black above each virus strain are geometric mean titers (GMT) of the reciprocal plasma dilution (FRNT50) resulting in 50% reduction in the number of infection foci. Numbers in red denote fold-change in GMT between virus strain on the left and the virus strain on the right of each panel. Red horizontal line denotes most concentrated plasma used. Samples were tested from n=19 participants, where n=6 were vaccinated only and n=13 were vaccinated and previously infected. p=0.006 for both (A) and (B) as determined by the Wilcoxon rank sum test.

**Table S1:**
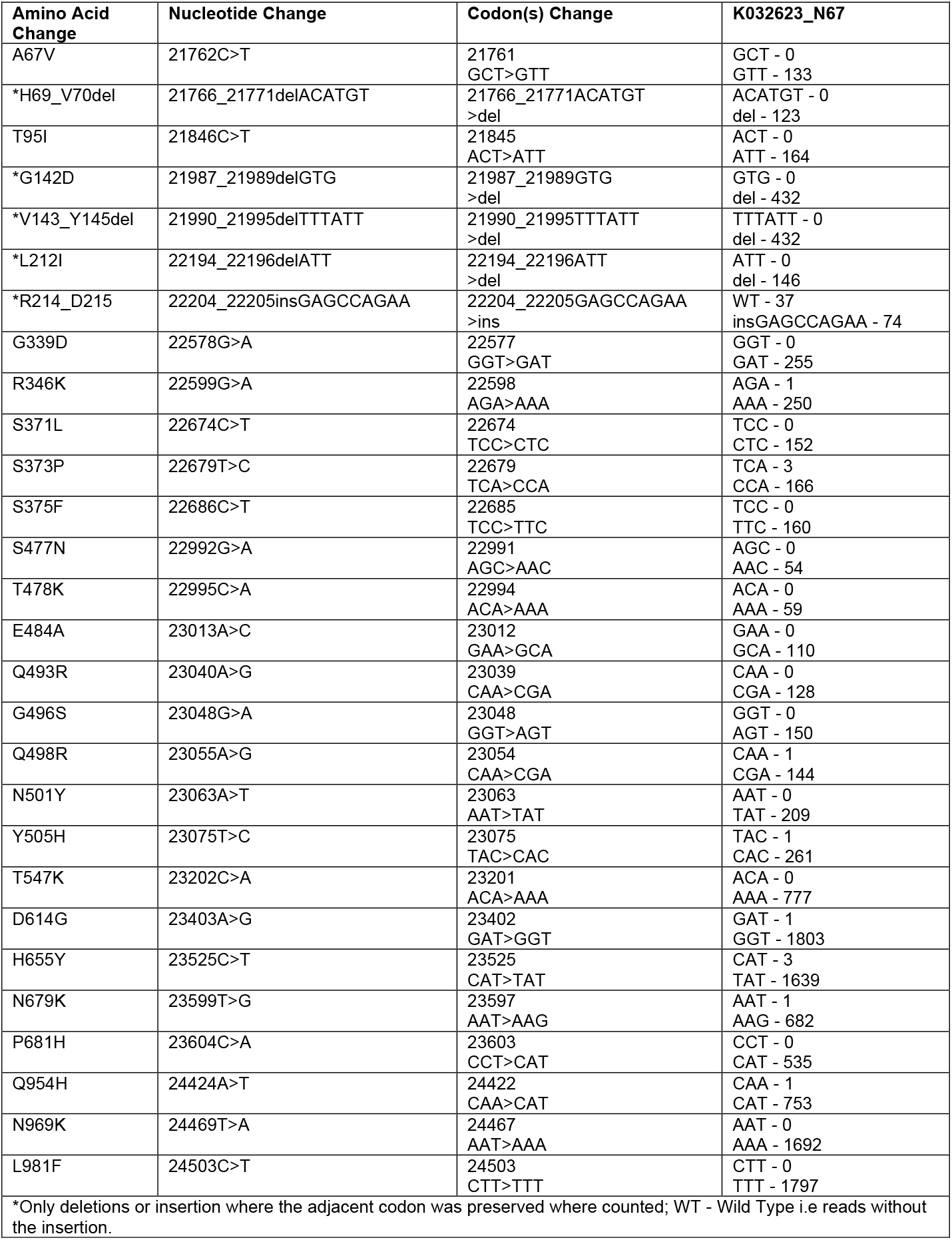
Codon frequency table This table shows the amino acid change, the nucleotide position of the genome, codon change and the frequency of the codon on the assembled genome.

| Table S1: Codon frequency table |  |  |  |
| --- | --- | --- | --- |
| This table shows the amino acid change, the nucleotide position of the genome, codon change and the frequency of the codon on the assembled genome. |  |  |  |
| Amino Acid Change | Nucleotide Change | Codon(s) Change | K032623_N67 |
| A67V | 21762C>T | 21761<br>GCT>GTT | GCT - 0<br>GTT - 133 |
| *H69_V70del | 21766_21771delACATGT | 21766_21771ACATGT<br>>del | ACATGT - 0<br>del - 123 |
| T95I | 21846C>T | 21845<br>ACT>ATT | ACT - 0<br>ATT - 164 |
| *G142D | 21987_21989delGTG | 21987_21989GTG<br>>del | GTG - 0<br>del - 432 |
| *V143_Y145del | 21990_21995delTTTATT | 21990_21995TTTATT<br>>del | TTTATT - 0<br>del - 432 |
| *L212I | 22194_22196delATT | 22194_22196ATT<br>>del | ATT - 0<br>del - 146 |
| *R214_D215 | 22204_22205insGAGCCAGAA | 22204_22205GAGCCAGAA<br>>ins | WT - 37<br>insGAGCCAGAA - 74 |
| G339D | 22578G>A | 22577<br>GGT>GAT | GGT - 0<br>GAT - 255 |
| R346K | 22599G>A | 22598<br>AGA>AAA | AGA - 1<br>AAA - 250 |
| S371L | 22674C>T | 22674<br>TCC>CTC | TCC - 0<br>CTC - 152 |
| S373P | 22679T>C | 22679<br>TCA>CCA | TCA - 3<br>CCA - 166 |
| S375F | 22686C>T | 22685<br>TCC>TTC | TCC - 0<br>TTC - 160 |
| S477N | 22992G>A | 22991<br>AGC>AAC | AGC - 0<br>AAC - 54 |
| T478K | 22995C>A | 22994<br>ACA>AAA | ACA - 0<br>AAA - 59 |
| E484A | 23013A>C | 23012<br>GAA>GCA | GAA - 0<br>GCA - 110 |
| Q493R | 23040A>G | 23039<br>CAA>CGA | CAA - 0<br>CGA - 128 |
| G496S | 23048G>A | 23048<br>GGT>AGT | GGT - 0<br>AGT - 150 |
| Q498R | 23055A>G | 23054<br>CAA>CGA | CAA - 1<br>CGA - 144 |
| N501Y | 23063A>T | 23063<br>AAT>TAT | AAT - 0<br>TAT - 209 |
| Y505H | 23075T>C | 23075<br>TAC>CAC | TAC - 1<br>CAC - 261 |
| T547K | 23202C>A | 23201<br>ACA>AAA | ACA - 0<br>AAA - 777 |
| D614G | 23403A>G | 23402<br>GAT>GGT | GAT - 1<br>GGT - 1803 |
| H655Y | 23525C>T | 23525<br>CAT>TAT | CAT - 3<br>TAT - 1639 |
| N679K | 23599T>G | 23597<br>AAT>AAG | AAT - 1<br>AAG - 682 |
| P681H | 23604C>A | 23603<br>CCT>CAT | CCT - 0<br>CAT - 535 |
| Q954H | 24424A>T | 24422<br>CAA>CAT | CAA - 1<br>CAT - 753 |
| N969K | 24469T>A | 24467<br>AAT>AAA | AAT - 0<br>AAA - 1692 |
| L981F | 24503C>T | 24503<br>CTT>TTT | CTT - 0<br>TTT - 1797 |
| *Only deletions or insertion where the adjacent codon was preserved where counted; WT - Wild Type i.e reads without the insertion. |  |  |  |

**Table S2:** Summary Table of Participants

|  | All | Vaccinated only | Infected and vaccinated |
| --- | --- | --- | --- |
| **Number of Participants** | 19 | 6 | 13 |
| **Age (years)** | 52 (39-67) | 54 (36-71) | 51 (45-63) |
| **Days post-vaccination** | 26 (14-33) | 14.5 (8.5-37.5) | 28 (18-32) |
| **Days post-infection** |  |  | 379 (127-468) |
| **Days post-infection to vaccination** |  |  | 353 (114-444) |
| **Date range of symptom onset** |  |  | Jun 2020 – Jul 2021 |
| **Male sex** | 7 | 2 | 5 |

All values are median (IQR) and inclusive of all samples used (early and late timepoints for 2 participants).

**Table S3:** Participant information per sample

| Sample | Participant | Age | Sex | Days post vaccination | Days between diagnostic swab and sample | Date symptom onset or diagnostic test | Infecting virus* |
| --- | --- | --- | --- | --- | --- | --- | --- |
| 1 | 1 | 60-69 | F | 10 | - | - | - |
| 2 | 2 | 70-79 | M | 10 | - | - | - |
| 3 | 2 | 70-79 | M | 45 | - | - | - |
| 4 | 3 | 30-39 | M | 14 | - | - | - |
| 5 | 4 | 70-79 | F | 10 | - | - | - |
| 6 | 4 | 70-79 | F | 48 | - | - | - |
| 7 | 5 | 30-39 | F | 10 | - | - | - |
| 8 | 6 | 30-39 | F | 33 | - | - | - |
| 9 | 7 | 40-49 | F | 14 | 458 | Jul-2020 | Ancestral |
| 10 | 8 | 60-69 | F | 63 | 468 | Jul-2020 | Ancestral |
| 11 | 9 | 20-29 | F | 31 | 487 | Aug-2020 | Ancestral |
| 12 | 10 | 20-29 | M | 37 | 493 | Jul-2020 | Ancestral |
| 13 | 11 | 60-69 | F | 28 | 378 | Jul-2020 | Ancestral |
| 14 | 12 | 60-69 | M | 26 | 379 | Jul-2020 | Ancestral |
| 15 | 13 | 40-49 | F | 32 | 479 | Aug-2020 | Ancestral |
| 16 | 14 | 50-59 | M | 30 | 370 | Sep-2020 | Ancestral |
| 17 | 15 | 40-49 | F | 22 | 456** | Jun-2020** | Ancestral/Delta |
| 18 | 16 | 40-49 | M | 18 | 83 | Jul-2021*** | Delta |
| 19 | 17 | 70-79 | M | 37 | 8 | Jul-2021 | Delta |
| 20 | 18 | 50-59 | F | 13 | 127 | Jul-2021*** | Delta |
| 21 | 19 | 60-69 | F | 14 | 103 | Jul-2021 | Delta |

\*Determined by infection wave in South Africa. First infection wave (April-October 2020) consisted of ancestral strains with the D614G mutation. Third infection wave (April-October 2021) was dominated by the Delta variant. \*\*Participant reinfected during Delta infection wave, sample is taken 3 months post-recovery of Delta infection. Asymptomatic during reinfection. \*\*\*Asymptomatic.

